# The integration of genetically-regulated transcriptomics and electronic health records highlights a pattern of medical outcomes related to increased hepatic *Transthyretin* expression

**DOI:** 10.1101/2021.07.14.21260525

**Authors:** Gita A. Pathak, Antonella De Lillo, Frank R. Wendt, Flavio De Angelis, Dora Koller, Brenda Cabrera Mendoza, Daniel Jacoby, Edward J. Miller, Joel N. Buxbaum, Renato Polimanti

**Affiliations:** Department of Psychiatry, Yale School of Medicine, West Haven, CT 06516, USA; VA CT Healthcare Center, West Haven, CT 06516, USA; Department of Biology, University of Rome Tor Vergata, Rome 00133, Italy; Section of Cardiovascular Medicine, Department of Internal Medicine, Yale School of Medicine, New Haven, CT 06510, USA; The Scripps Research Institute, La Jolla, CA 92037, USA

**Keywords:** gene expression, TTR, amyloidosis, retinol, thyroxine, UK Biobank, phenome-wide association study

## Abstract

**Background:** Transthyretin (TTR) is a multi-function protein involved in the systemic transport of retinol and thyroxine. It also participates in the neuronal response to stress and proteolysis of few specific substrates. TTR is also the precursor of the fibrils that compromise organ function in the familial and sporadic systemic amyloidoses (ATTR). RNA-interference and anti-sense therapeutics targeting *TTR* hepatic transcription have been shown to reduce TTR amyloid formation. The goal of our study was to investigate the role of genetic regulation of *TTR* transcriptomic variation in human traits and diseases.

**Methods and Findings:** We leveraged genetic and phenotypic information from the UK Biobank and transcriptomic profiles from the GTEx (Genotype-Tissue Expression) project to test the association of genetically regulated *TTR* gene expression with 7,149 traits assessed in 420,531 individuals. We conducted a joint multi-tissue analysis of *TTR* transcription regulation and identified an association with a specific operational procedure related to secondary open reduction of fracture of bone (p=5.46×10^−6^, false discovery rate q=0.039). Using tissue-specific *TTR cis* expression quantitative trait loci, we demonstrated that the association is driven by the genetic regulation of *TTR* hepatic expression (odds ratio [OR] = 3.46, 95% confidence interval [CI] = 1.85-6.44, p = 9.51×10^−5^). Although there is an established relationship of retinol and thyroxine abnormalities with bone loss and the risk of bone fracture, this is the first evidence of a possible effect of *TTR* transcriptomic regulation. Investigating the UK Biobank electronic health records available, we investigated the comorbidities affecting individuals undergoing the specific surgical procedure. Excluding medical codes related to bone fracture events, we identified a pattern of health outcomes that have been previously associated with ATTR manifestations. These included osteoarthritis (OR=3.18, 95%CI=1.93-4.25, p=9.18×10^−8^), carpal tunnel syndrome (OR=2.15, 95%CI=1.33-3.48, p=0.002), and a history of gastrointestinal diseases (OR=2.01, 95%CI=1.33-3.01, p=8.07×10^−4^).

**Conclusions:** The present study supports the notion that *TTR* hepatic expression can affect health outcomes linked to physiological and pathological processes presumably related to the encoded protein. Our findings highlight how the integration of omics information and electronic health records can successfully dissect the complexity of multi-function proteins such as TTR.

## Background

Transthyretin (TTR) is a multi-function protein involved in a range of physiological processes [1, 2]. The bulk of circulating TTR is produced in the liver, but its synthesis has been reported in other tissues (e.g., choroid plexus neurons under stress, retinal epithelial cells, Schwann cells, pancreatic epithelium, and skin) [1, 2]. The TTR tetramer binds thyroxine (T4) and retinol (bound to the retinol-binding protein 4, holo-RBP4) and plays a role in their transportation through the circulatory system and into the central nervous system [1, 2]. TTR knockout mouse models showed a consistent reduction of T4 (50%) and holo-RBP4 (95%) in blood [3, 4]. This suggests that TTR regulation may impact health outcomes linked to retinol and thyroid homeostasis. In addition to its transport function, TTR is a metallopeptidase with three recognized substrates: apolipoprotein A-I (apoA-I), neuropeptide Y (NPY), and amyloid-beta peptide (Aβ) [1]. TTR can cleave the C-terminus of apoA-I, decreasing cholesterol efflux and potentially affecting lipid metabolism and the development of atherosclerosis [5]. In vitro TTR has been shown to cleave different forms of Aβ, but many studies have shown that it can prevent the formation of new aggregates, oligomers and fibrils with little or no evidence that proteolysis is required for its anti-Aβ role in vitro or in vivo [6].

TTR is best known for its role as a fibril precursor involved in the pathogenesis of human systemic amyloidoses (e.g., familial amyloidotic polyneuropathy, familial amyloid cardiomyopathy) [7]. TTR amyloidogenesis is initiated by the dissociation of the tetramer into monomers, which is followed by misfolding and aggregation into non-native oligomers, and finally the highly structured amyloid fibrils [8]. The accumulation of the TTR amyloid fibrils in multiple organs causes different symptoms depending on the site of the fibrils deposition. TTR-associated amyloidosis (ATTR) can be familial or sporadic. The familial forms of ATTR (ATTRm) are caused by coding mutations in the *TTR* gene with an autosomal dominant inheritance [9]. ATTRm patients have multiple organ involvement with peripheral and autonomic neuropathy, cardiomyopathy, gastrointestinal impairment, nephropathy, and ocular deposition with vitreous opacities. Wild-type TTR (ATTRwt; i.e. TTR protein without amyloidogenic mutations) can also form amyloid fibrils depositing primarily in the heart [10]. Although ATTRwt is generally associated with late-onset cardiomyopathy, there are other sites of deposition which may be associated with clinical symptoms, most prominently carpal tunnel syndrome and lumbar spinal stenosis [11]. Gastrointestinal deposits of TTRwt are also common in the elderly, being found in 8-20% of gastrointestinal biopsies in such individuals [12]. In addition, many patients undergoing hip and knee replacements have ATTRwt deposition in the excised abnormal joints [13].

Due to their clinical heterogeneity, ATTRm and ATTRwt can be often misdiagnosed as other, more common disorders which present similar symptomatology. The diagnosis of ATTRm is frequently only made many years after the onset of symptoms [14]. With respect to ATTRwt, its prevalence has increased in recent years, largely because of heightened awareness in the medical community and improved diagnostic tools, but the disorder is still underdiagnosed [15]. Additionally, it is now known that ATTRwt can be identified in individuals less than 50 years of age, challenging the concept that this is a disease exclusively of the elderly [11]. Although *TTR* coding mutations are the cause of ATTRm, other factors contribute to the complex genotype-phenotype correlation (the same point mutation may be associated with different phenotypic combinations even within the same family) [16]. Non-coding variants regulating *TTR* transcription have been suggested to modify the phenotypic presentation in carriers of amyloidogenic mutations [17-19]. Similarly, transcriptomic regulation appears to have a role in the onset of ATTRwt in non-carriers [20, 21]. Other non-coding regulatory mechanisms such as epigenetic changes seem to affect ATTR genotype-phenotype correlation [22, 23].

Since organ involvement in ATTR pathogenesis requires an ongoing supply of mutant TTR precursor, RNA-interference (RNAi) and anti-sense therapeutics capable of silencing *TTR* gene expression in the liver have been shown to decrease total TTR production by as much as 80%, with subsequent reduction of symptoms associated with the deposits [24-26]. Recently, CRISPR-Cas9 in vivo gene editing targeting *TTR* gene expression in liver showed a reduction of TTR protein hepatic production in ATTRm patients [27]. To explore the relationship between physiological and pathological processes linked to *TTR* transcriptomic regulation, we conducted a phenome-wide investigation, applying transcriptome prediction models to 7,149 traits assessed in up to 420,531 individuals and subsequently investigated *TTR* tissue-specific contributions using a Mendelian randomization (MR) framework.

## Methods

### Study Design

Leveraging genome-wide and transcriptomic data, we tested the effect of *TTR* transcriptomic variation across the human phenotypic spectrum. Specifically, we estimated the joint effect sizes of genetic variants for *TTR* gene expression. This permitted us to evaluate the genetically-regulated transcriptomic association of *TTR* with human traits and diseases, independent of the transcriptomic changes induced by the environment and the phenotypes investigated [28]. Subsequently, we conducted a two-sample *cis*-MR analysis to assess the contributions of the tissue-specific gene expression and investigated electronic health records (EHR) of UK Biobank (UKB) participants to identify the spectrum of medical outcomes related to *TTR* gene.

### Datasets

The UKB cohort is an open-access resource available to investigate a wide range of severe and life-threatening illnesses as well as normal-range traits [29]. This initiative has recruited more than 500,000 people (54% females; age at recruitment: 37-73 years) collecting information on their diet, cognitive function, work history, health status, and other relevant phenotypes. More than 7,000 phenotypic traits are available to be investigated in this cohort so far (Supplementary Table 1). Leveraging UKB genome-wide association statistics, we derived information regarding genetic variants regulating *TTR* gene expression and data related to the health and disease status of its participants. These data are available at https://pan.ukbb.broadinstitute.org/downloads. A detailed description of the methods used to generate these data is available at https://pan.ukbb.broadinstitute.org/. Briefly, the genome-wide association analysis was conducted using the SAIGE (Scalable and Accurate Implementation of GEneralized) mixed model [30] and including a kinship matrix as a random effect and covariates as fixed effects. The covariates included age, sex, age×sex, age^2^, age^2^×sex, and the top-10 within-ancestry principal components. We investigated association statics derived from UKB participants of European descent (N=420,531), because no large-scale multi-tissue transcriptomic datasets are available to investigate other ancestries.

Information regarding the genetic regulation of *TTR* gene expression across multiple tissues was derived from the Genotype-Tissue Expression project (GTEx) [31]. This is a comprehensive public resource to study tissue-specific gene expression and its regulation. Samples were collected from 54 non-diseased tissue sites across nearly 1000 individuals, primarily for molecular assays including whole-genome sequencing, whole-exome sequencing, and RNA sequencing (RNA-seq). We used the GTEx V8 [32], which includes 17,382 RNA-Seq samples from 948 donors (312 women and 636 men). Supplementary Figure 1 shows *TTR* transcriptomic profiles in the 54 human tissues available in GTEx V8. The *cis*-regulatory effect of transcriptomic variation was assessed investigating the association of genetic variants located within 1 Mb up- and downstream of the transcription start site (for *TTR* gene, hg19 chr18:28,171,730-30,178,986). A detailed description of the GTEx methods (i.e., preprocessing, expression quantification, and association analysis) used is available at https://gtexportal.org/.

Briefly, the *cis* expression quantitative locus (eQTL) mapping was conducted using FastQTL and considering as covariate the top-five genotyping principal components, PEER (Probabilistic Estimation of Expression Residuals) factors, sequencing platform, sequencing protocol, and sex. The eQTL mapping was conducted in 49 GTEx tissues that were assessed in at least 70 samples.

### Gene Expression Prediction from TTR Association Statistics

To evaluate the genetically-regulated *TTR* transcriptomic variation, we used pre-trained prediction models available in S-PrediXcan method [33]. These gene expression weights were derived from GTEx v8 transcriptomic data using the fine-mapping software DAP-G [34] with a biologically informed prior, Multivariate Adaptive Shrinkage in R (MASHR) [35]. With respect to *TTR* transcriptomic variation, MASHR-based models were available for eight GTEx tissues: adrenal gland, aorta artery, nucleus accumbens (basal ganglia), putamen (basal ganglia), esophagus (gastroesophageal junction), minor salivary gland, skin (lower leg), and whole blood. These models were tested with respect to *TTR* association statistics available from the UKB analysis using S-PrediXcan [33]. Since the MASHR-based models were derived from samples collected from individuals of European descent, we used UKB association statistics calculated from UKB participants of European descent (N=420,531) to avoid population stratification biases. To boost the statistical power, we combined tissue-specific information using S-MultiXcan, which permitted us to perform a joint multi-tissue analysis accounting for transcriptomic correlation across the tissues tested [36]. S-MultiXcan analysis of *TTR* gene was conducted across 7,149 traits (Supplementary Table 1). A false discovery rate correction (FDR q<0.05) was applied to account for the number of traits tested.

### Two-sample Mendelian Randomization

The results obtained from the S-MultiXcan analysis were investigated further using a two-sample MR analysis. This approach permitted us to estimate the putative causal effect of the tissue-specific genetically-regulated *TTR* transcriptomic variation on the traits of interest. To maximize the statistical power, we included all linkage disequilibrium (LD) independent *TTR cis* eQTL in genetic instruments derived from the 49 GTEx tissues available. For each tissue, we conducted a clumping considering an LD cutoff of R^2^ =0.001 within a 10,000-kilobase window and using the 1,000 Genomes Project Phase 3 reference panel for European populations. Since we considered LD-independent *TTR cis* eQTL independently from their individual statistical significance, we conducted an MR analysis using the robust adjusted profile score (MR-RAPS) approach [37]. This is a method specifically designed to conduct causal inference analysis based on weak genetic instruments, accounting for the widespread pleiotropy of complex traits.

### Co-morbidity Analysis using UKB Electronic Health Records

To investigate the comorbidities of the traits identified as associated with genetically determined *TTR* transcriptomic variation through S-MultiXcan and MR-RAPS analyses, we used EHRs available from UKB. Specifically, we evaluated the occurrence of ICD-10 (International Classification of Disease, 10^th^ revision) and OPCS-4 (Office of Population Censuses and Surveys Classification of Interventions and Procedures, 4^th^ revision) codes between cases presenting S-MultiXcan/MR-RAPS-identified traits and controls matched by ancestry, age, sex, Townsend deprivation index, and UKB recruiting center. Based on the matching criteria, we maximized the number of control for each case to boost the statistical power of our analysis.

Townsend deprivation index is a measure of material deprivation incorporating unemployment, non-car ownership, and non-home ownership [38]. At recruitment, each UKB participant is assigned a deprivation score corresponding to their address location Comparing the occurrence of ICD-10 and OPCS-4 codes between cases *vs*. matched controls, we calculated odds ratios (OR) and the corresponding 95% confidence intervals (95%CI) and applied an FDR correction (q<0.05) accounting for the number of medical outcomes tested.

## Results

Leveraging MASHR-based models and the S-MultiXcan approach, we conducted a phenome-wide analysis across 7,149 traits and identified one association surviving FDR correction (q < 0.05; Figure 1): a specific operational procedure recorded in hospital inpatient records (Phenotype ID 41200-W231; OPCS-4: W23.1 – Secondary open reduction of fracture of bone and intramedullary fixation; p=5.46×10^−6^, FDR q=0.039). To confirm this finding, we applied the MR-RAPS approach to estimate the effect of the tissue-specific transcriptomic regulation of *TTR* gene on the odds of presenting the phenotype 41200-W231 among UKB participants. In liver, one standard deviation increase in the genetically-regulated transcriptomic profile of *TTR* gene was associated with a 3.46-fold increase in the odds of presenting the phenotype 41200-W231 (95%CI=1.85-6.44, p=9.51×10^−5^ ; Figure 2). Among the other tissues, we observed the second strongest effect in the pancreas (OR=2.83, 95%CI=1.2-6.69, p=0.017). This is in line with the *TTR* expression where liver and pancreas present the highest levels among the tissues available from GTEx (median transcripts per million: 2,734 and 202.8, respectively; see GTEx data in Supplementary Figure 1). Because *TTR* gene is mainly expressed in the liver, we conducted a phenome-wide analysis to identify health outcomes not detected by the MultiXcan analysis. We identified a small negative effect of hepatic *TTR* expression on phenotype 1697 (“comparative height size at age 10”; beta=-0.024, p=5.15×10^−5^). In the MultiXcan analysis, this phenotype was only nominally associated with the genetically-regulated transcriptomic profile of *TTR* gene (S-MultiXcan p=0.008). Beyond these two traits, we found limited concordance between these approaches (Supplementary Figure 2).

**Figure 1:**
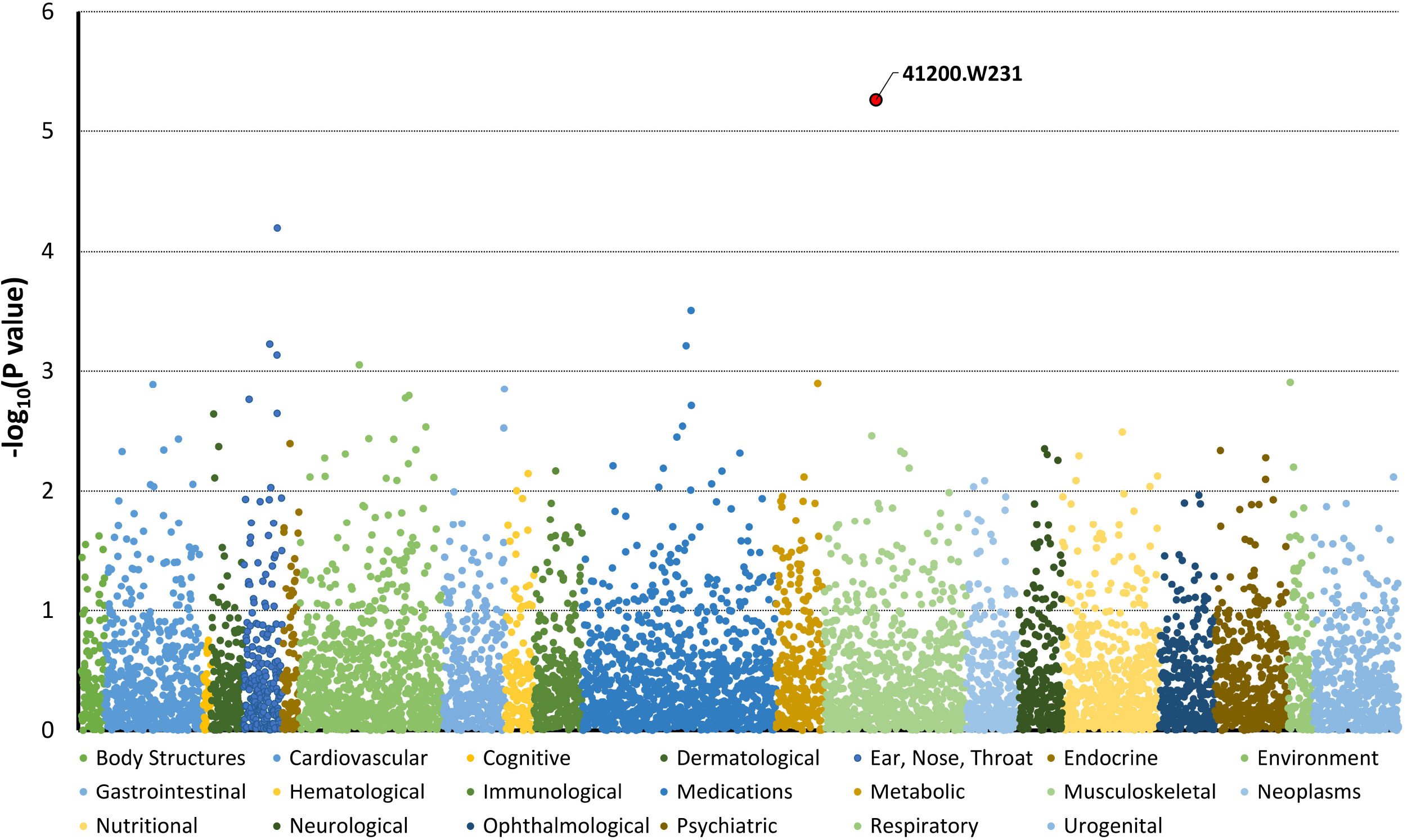
Phenome-wide association statistics of the joint multi-tissue analysis of the genetically regulated *TTR* transcriptomic variation across the 7,149 traits tested.

**Figure 2:**
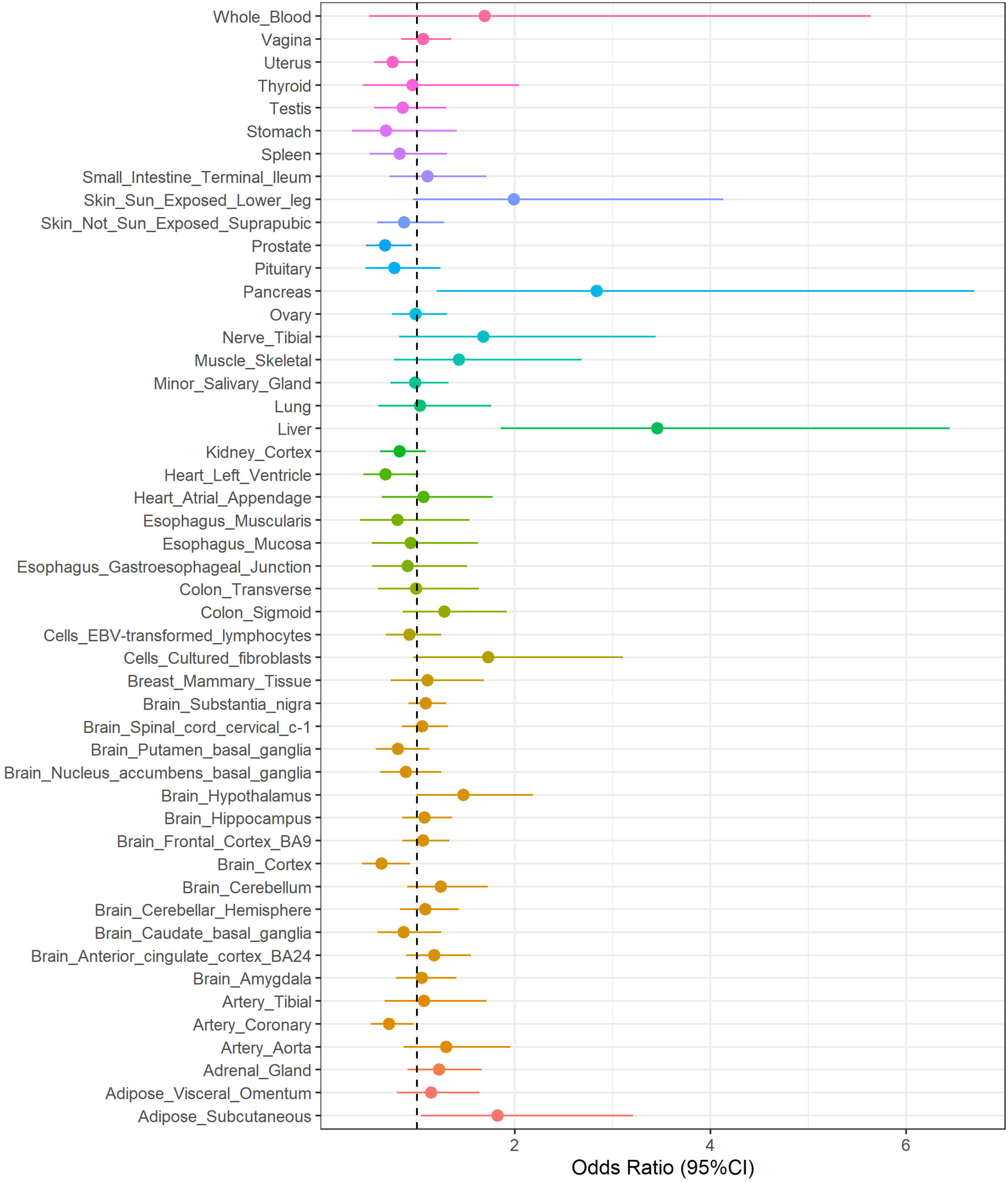
Tissue-specific effect of genetically regulated *TTR* transcriptomic variation on the 41200-W231 phenotype calculated via an MR analysis using the robust adjusted profile score.

We observed a large effect of the genetically-regulated *TTR* transcriptomic variation in the liver with the phenotype 41200-W231 (OR=3.46; 95%CI=1.85-6.44) that is reported only for 290 UKB participants of European descent (0.07%). None of them is a carrier of one of the *TTR* coding mutations defined as amyloidogenic in the curated list available at http://www.amyloidosismutations.com/. We verified whether the strong case-control imbalance of the phenotype 41200-W231 affects the association statistics. Conducting a MultiXcan analysis across 22,196 genes, we confirmed that *TTR* is the gene with the strongest association and that there is no inflation in the transcriptome-wide association statistics (λgc=0.996; Supplemental Figure 3). To investigate the comorbidities of 41200-W231 cases, we verified whether this group of individuals present phenotypic presentations linked to physiological and pathological processes that may be affected by a genetic predisposition to a high hepatic expression of *TTR* gene. To improve power of detection for 41200-W231 comorbidities, we tested ICD-10 and OPCS-4 codes that were present in at least 5% of the 41200-W231 cases. Comparing 41200-W231 cases with 25,229 controls matched by ancestry, age, sex, Townsend deprivation index, and UKB recruiting center (Supplementary Table 2; controls did not include carriers of *TTR* amyloidogenic mutations), we identified several associated medical outcomes after applying an FDR multiple testing correction (q<0.05; Supplementary Tables 3 and 4). Excluding outcomes that can be directly reconducted to 41200-W231 phenotype (e.g., ICD-10 codes related to injuries and factors influencing contact with health services), we observed that 41200-W231 cases present a pattern of medical outcomes that may be due to hepatic *TTR* transcriptomic regulation (Table 1). The strongest 41200-W231 association was with osteoarthritis (ICD-10: M19.9, OR=3.18, 95%CI=1.93-4.25, p=9.18×10^−8^). Considering ICD-10 and OPCS-4 codes, we observed a convergence related to medical outcomes involving gastrointestinal system (ICD-10: Z87.1 “history of gastrointestinal diseases”, OR=2.01, 95%CI=1.33-3.01, p=8.07×10^−4^ ; OPCS-4: G45.9 “Unspecified diagnostic fiberoptic endoscopic examination of upper gastrointestinal tract”, OR=1.89, 95%CI=1.37-2.61 p=9.75×10^−5^) and carpal tunnel (ICD-10: G56.0 “Carpal tunnel syndrome”, OR=2.15, 95%CI=1.33-3.48, p=0.002; OPCS-4: A65.1 “Carpal tunnel release”, OR=2.2, 95%CI=1.36-3.56, p=0.001). In addition to outcomes related to physical health, we also observed associations with traits related to mental health: depressive episode (ICD-10: F32.9; OR=2.86, 95%CI=1.93-4.25, p=1.9×10^−7^), personal history of psychoactive substance abuse (ICD-10: Z86.4; OR=2, 95%CI=1.46-2.73, p=3.5×10^−5^), tobacco use (ICD-10: Z72.0, OR=2.46, 95%CI=1.45-4.15, p=5.51×10^−4^), and harmful nicotine use (ICD-10: F17.1, OR=2.13, 95%CI=1.32-3.44, p=0.002).

**Table 1:** Significant association of 41200.W231 status with diagnostic and operational codes (FDR q<0.05). Outcomes directly related to 41200.W231 were not reported. Full results are listed in Supplementary Table 3 and 4. Abbreviations: International Classification of Disease, 10th revision (ICD-10); Office of Population Censuses and Surveys Classification of Interventions and Procedures, 4th revision (OPCS-4); Odds ratio (OR); Confidence interval (CI); Not Elsewhere Classified (NEC).

|  | Code | 41200.W231 cases (%) | matched controls (%) | OR | 95% CI |
| --- | --- | --- | --- | --- | --- |
| ICD-10 | M19.9 Arthrosis, unspecified | 0.083 | 0.028 | 3.18 | 2.08-4.86 |
|  | F32.9 Depressive episode, unspecified | 0.097 | 0.036 | 2.86 | 1.93-4.25 |
|  | E11.9 Without complications | 0.117 | 0.049 | 2.57 | 1.79-3.69 |
|  | M81.99 Osteoporosis, unspecified (Site unspecified) | 0.052 | 0.014 | 3.93 | 2.31-6.68 |
|  | N17.9 Acute renal failure, unspecified | 0.059 | 0.017 | 3.63 | 2.20-5.98 |
|  | J45.9 Asthma, unspecified | 0.159 | 0.08 | 2.18 | 1.58-3.00 |
|  | Z86.4 Personal history of psychoactive substance abuse | 0.166 | 0.09 | 2 | 1.46-2.73 |
|  | D64.9 Anemia, unspecified | 0.076 | 0.035 | 2.28 | 1.47-3.54 |
|  | R11 Nausea and vomiting | 0.079 | 0.037 | 2.22 | 1.44-3.42 |
|  | I10 Essential (primary) hypertension | 0.321 | 0.233 | 1.56 | 1.22-2.00 |
|  | Z72.0 Tobacco use | 0.055 | 0.023 | 2.46 | 1.48-4.10 |
|  | Z87.1 Personal history of diseases of the digestive system | 0.09 | 0.047 | 2.01 | 1.34-3.02 |
|  | J44.9 Chronic obstructive pulmonary disease, unspecified | 0.052 | 0.022 | 2.45 | 1.45-4.15 |
|  | G56.0 Carpal tunnel syndrome | 0.062 | 0.03 | 2.15 | 1.32-3.49 |
|  | E78.0 Pure hypercholesterolemia | 0.152 | 0.097 | 1.66 | 1.20-2.29 |
|  | F17.1 Harmful use | 0.062 | 0.03 | 2.13 | 1.32-3.44 |
|  | Z88.0 Personal history of allergy to penicillin | 0.093 | 0.052 | 1.87 | 1.26-2.78 |
|  | K52.9 Non-infective gastro-enteritis and colitis, unspecified | 0.076 | 0.041 | 1.91 | 1.23-2.97 |
|  | N39.0 Urinary tract infection, site not specified | 0.072 | 0.04 | 1.9 | 1.21-2.97 |
|  | E66.9 Obesity, unspecified | 0.069 | 0.038 | 1.9 | 1.20-3.00 |
|  | I20.9 Angina pectoris, unspecified | 0.076 | 0.043 | 1.84 | 1.18-2.87 |
|  | R69 Unknown and unspecified causes of morbidity | 0.083 | 0.048 | 1.79 | 1.17-2.73 |
|  | K62.5 Hemorrhage of anus and rectum | 0.059 | 0.033 | 1.82 | 1.11-2.99 |
|  | K59.0 Constipation | 0.059 | 0.034 | 1.75 | 1.07-2.87 |
|  | I25.9 Chronic ischemic heart disease, unspecified | 0.055 | 0.032 | 1.76 | 1.06-2.92 |
|  | K21.0 Gastro-esophageal reflux disease with esophagitis | 0.052 | 0.03 | 1.77 | 1.04-3.00 |
|  | R19.4 Change in bowel habit | 0.062 | 0.038 | 1.66 | 1.02-2.69 |
| OPCS-4 | G45.9 Unspecified diagnostic fiberoptic endoscopic examination of upper gastrointestinal tract | 0.155 | 0.088 | 1.89 | 1.37-2.60 |
|  | A65.1 Carpal tunnel release | 0.062 | 0.029 | 2.2 | 1.38-3.52 |
|  | Z92.6 Abdomen NEC | 0.062 | 0.035 | 1.82 | 1.13-2.93 |
|  | J18.3 Total cholecystectomy NEC | 0.066 | 0.039 | 1.72 | 1.08-2.75 |
|  | Z28.2 Caecum | 0.114 | 0.078 | 1.52 | 1.05-2.19 |
|  | Z27.4 Duodenum | 0.141 | 0.102 | 1.45 | 1.04-2.02 |

## Discussion

Leveraging genomic and transcriptomic datasets, we have demonstrated that increased hepatic *TTR* transcription due to genetic regulation is linked to a set of clinical phenotypes. We hypothesize that the effect of genetically-regulated *TTR* transcriptomic variation on TTR protein production. RNAi and anti-sense therapeutics targeting TTR transcriptomic regulation in liver showed a reduction in the hepatic production of TTR protein [24-26]. Recently, an in vivo gene-editing therapeutic agent has been shown to induce an 87% reduction of serum TTR protein concentration by altering the *TTR* gene sequence in liver (i.e., targeted knockout) to reduce its hepatic transcription [27]. Accordingly, the association between genetically-regulated *TTR* transcriptomic variation and certain health outcomes can be due to two mechanisms: i) direct effect – increased TTR production with the protein being toxic (as in amyloid); ii) altered homeostasis of molecules carried by TTR (e.g., T4).

In our phenome-wide analysis, genetically-regulated *TTR* transcriptomic variation was positively associated with increased odds of having a secondary open reduction of fracture of bone and intramedullary fixation (phenotype: 41200-W231; OPCS-4: W23.1). As mentioned, this effect is due to the transcriptomic variation regulated by genetic variation and not transcriptomic changes induced by environmental factors or physiological and pathological conditions of the individuals investigated. Additionally, the association was consistent across two analytic approaches based on different genetic estimators of *TTR* gene expression (i.e., joint-tissue gene expression modeling and tissue-specific *cis*-regulatory mediation). In line with *TTR* tissue distribution, hepatic gene regulation was the primary driver of this effect. As expected, individuals having W23.1 code in their OPCS-4 records also report ICD-10 codes related to bone fractures and fallings (from slipping, tripping, and stumbling) on same level and on/from stairs and steps (Supplementary Table 3). The susceptibility to bone fractures requiring open surgeries can be linked to the role of TTR protein in retinol and T4 transport. Altered retinol homeostasis has been associated with an increased risk of hip fractures [39-41]. In mice, excess vitamin A intake (high dosages for a short period of time and clinically relevant dosages over prolonged times) has been associated with decreased cortical bone thickness and increased fracture risk [42]. There is also a well-established relationship between T4 levels and the risk of bone fracture [43]. The mechanism is related to the effect of T4 on adult bone turnover and maintenance [44]. This effect is particularly evident in both hyperthyroidism and hypothyroidism. In hyperthyroidism, individuals have an excessive amount of the hormone T4, causing bone loss among several other symptoms [44]. In hypothyroidism, individuals are usually treated with levothyroxine (a synthetic form of T4) that can lead to a bone density reduction as a side effect [45]. As demonstrated by knockout models, TTR plays an important role in maintaining retinol and T4 homeostasis [3, 4]. Accordingly, individuals predisposed to having high hepatic *TTR* expression could have an increased risk of bone fracture and subsequent corrective surgeries due to the altered blood levels of retinol and/or T4. The same mechanism could also explain the association of the genetically-regulated *TTR* expression in the liver with the likelihood of reporting to be shorter than average at 10 years of age (phenotype 1697 “comparative height size at age 10”). In particular, TTR could affect the regulatory effect of T4 on endochondral ossification, which is essential for skeletal development and linear growth [44].

Since the W23.1 code is present in the OPCS-4 records of 290 out of the 426,884 UKB participants investigated in the present study, we decided to verify whether these individuals present co-morbidities that cannot be attributed directly to a bone fracture and its subsequent surgeries. Comparing 41200-W231 cases with 25,229 matched controls, we identified a pattern of medical outcomes. The strongest evidence was observed for osteoarthritis (ICD-10: M19.9). TTR amyloid deposits have been reported in the articular cartilage of patients affected by osteoarthritis, suggesting a role in the pathogenesis of the disease [46]. 41200-W231 cases have also increased odds of having codes for carpal tunnel syndrome (ICD-10: G56.0) and carpal tunnel release (OPCS-4: A65.1) in their health records. Carpal tunnel syndrome is a recognized early sign of ATTR [47]. In the general population, carpal tunnel release has a lifetime prevalence of 3.1% [48]. This is in line with its frequency in our matched controls (2.9%) while it was 6.2% in 41200-W231 cases. TTR amyloid deposition has been documented in the transverse carpal ligament in between 10 and 35% of patients undergoing carpal tunnel release and in as many as 60% of cases with *TTR* amyloidogenic mutations [49]. Gastrointestinal symptoms were also supported by concordant evidence from ICD-10 and OPCS-4 codes. Compared to matched controls, 41200-W231 cases showed higher odds of presenting codes related to diseases of the digestive system (ICD-10: Z87.1), nausea (ICD-10: R11), non-infective gastro-enteritis (ICD-10: K52.9), Constipation (ICD-10: K59.0), gastro-esophageal reflux disease (ICD-10: K21.0), change in bowel habit (ICD-10: R19.4), endoscopic examination of the upper gastrointestinal tract (OPCS-4: G45.9), and different types of gastrointestinal tract surgery (OPCS-4: Z92.6, Z28.2, and Z27.4). In the Transthyretin Amyloidosis Outcomes Survey (THAOS), gastrointestinal symptoms were reported in 59% of the patients enrolled, indicating that this is a common manifestation for ATTR [50]. Other 41200-W231-associated health outcomes also overlap with known ATTR symptoms including renal failure (ICD-10: N17.9) [51], urinary tract infection (ICD-10: N39.0) [52], and angina pectoris (ICD-10: I20.9) [53]. Beyond the medical codes that can be linked to ATTR symptoms, we observed certain outcomes that may be linked to other functions of TTR protein. In particular, codes related to mental health (ICD-10: F32.9 depressive episode, Z86.4 psychoactive substance abuse, and F17.1 harmful nicotine use) may be linked to the role of TTR in the physiology of the nervous system. In knockout mice, TTR showed multiple effects on the central nervous system [2]. In particular, the absence of TTR was associated with a reduction in depressive-like behavior and an increase in exploratory activity [54]. In human subjects, low cerebrospinal fluid TTR levels were associated with major depression, suicidal ideation, and low serotonin function [55]. In our study, we focused on *TTR* hepatic expression, observing a possible link with depressive symptoms and substance use. Further studies focused on choroid plexus TTR synthesis will be needed to investigate the potential role of TTR in mental health.

In summary, our results support that the hepatic expression of *TTR* gene can affect several health outcomes due to its impact on physiological and pathological processes. With respect to the TTR-related physiological processes, we provide evidence that variation in *TTR* gene regulation can lead to medical outcomes due to the altered homeostasis of retinol and T4. Beyond the role of TTR in ligand transport, our findings highlight that genetically regulated *TTR* transcriptomic profile could be related to certain psychiatric traits in line with previous evidence about TTR function in brain physiology.

Although we demonstrate that the combination of genomic information and electronic health records may expose previously unsuspected TTR-associated diseases manifestations, there is still much to learn about the hepatic regulation of TTR transcription and its relationship to the disorders associated with TTR deposition as well as those in which at present the relationship is unexpected. We also acknowledge several limitations. While our main finding (i.e., the association of genetically-regulated *TTR* expression with 41200-W231 phenotype) was confirmed by two independent methods (i.e., MultiXcan and MR-RAPS), there was a limited convergence between these two approaches (Supplementary Figure 2). This is most likely because the current transcriptomic datasets are underpowered to investigate *TTR* gene regulation. Larger datasets will be needed to conduct more powerful studies of genetic regulation of *TTR* transcriptomic variation. Although multiple studies have shown that suppression of *TTR* hepatic transcription leads to a reduction in the concentration of TTR protein in the circulation [24-27], our study lacks information related to the amount of TTR protein synthesized and secreted in response to the increase in hepatic transcription. Hence, it is unclear whether the increased transcription results in an increase in amyloid precursor available for aggregation with subsequent tissue compromising fibrillogenesis. Similarly, we do not have data from the subjects regarding the circulating levels of the TTR ligands (i.e., T4 and holo-RBP4), abnormalities of which have been associated with susceptibility to fracture. Hence, we cannot ascertain the molecular mechanism relating increased hepatic transcription to identified clinical outcomes. Finally, our comorbidity analysis of 41200-W231 cases showed evidence consistent with symptoms that can be related to the pathologic effects of the TTR protein. However, we do not have information whether the symptoms observed in 41200-W231 cases are due to TTR amyloid deposits. We investigated only UKB participants of European descent, because no large-scale genomic and transcriptomic datasets are available to investigate other ancestries. This represents a major limitation for our study and for the entire field. Indeed, as other biomedical areas, certain minority groups are under-investigated in ATTR studies [56]. Acknowledging these limitations, our findings should be considered a proof of concept regarding the integration of omics information and EHRs to investigate the complexity of multi-function proteins such as TTR suggesting further studies leading to greater knowledge of their involvement in human diseases.

## Data Availability

All data discussed in this study are provided in the article and in the Supplementary Material.

## Declarations

### Ethics approval and consent to participate

Owing to the use of previously collected, deidentified data, this study did not require institutional review board approval.

### Consent for publication

Not Applicable.

### Availability of data and materials

All data discussed in this study are provided in the article and in the Supplementary Material.

## Authors’ contributions

RP designed the study. GAP, ADL, and RP conducted the statistical analysis and drafted the manuscript. GAP, ADL FRW, FDA, DK, BCM, RP participated in the data acquisition and analysis. All authors participated in interpretation of data and critical revision of the manuscript for important intellectual content. RP obtained the funding for this study. All the authors revised and approved the final version of this manuscript.

## Acknowledgements

This research has been conducted using the UK Biobank Resource (application reference no. 58146). We thank the research groups contributing to the GTEx Project and the Pan-UKB analysis for making their data publicly available.

