## Supplementary Figures for "The integration of genetically-regulated transcriptomics and electronic health records highlights a pattern of medical outcomes related to increased hepatic *Transthyretin* expression"

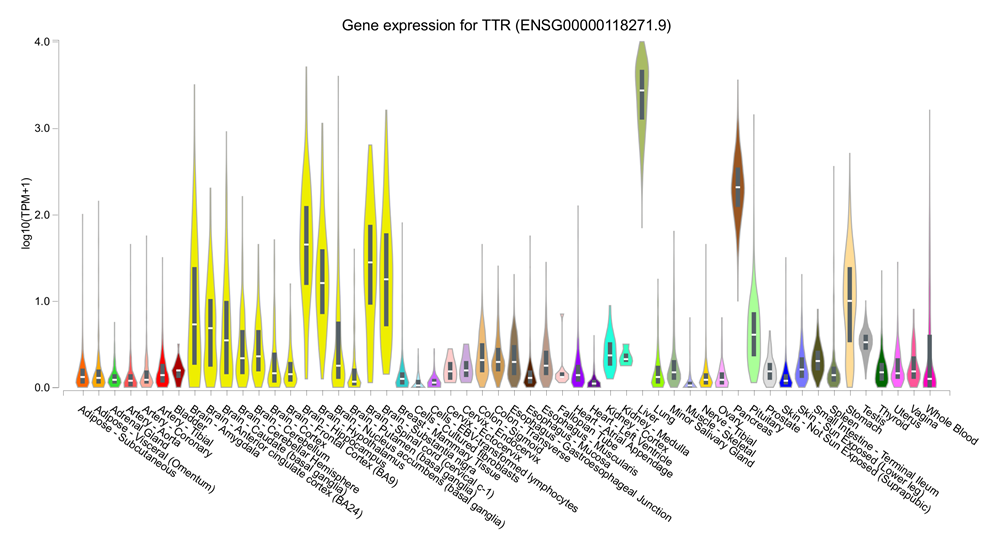

**Supplementary Figure 1**: *TTR* transcriptomic profile in 54 human tissues available from GTEx V8.

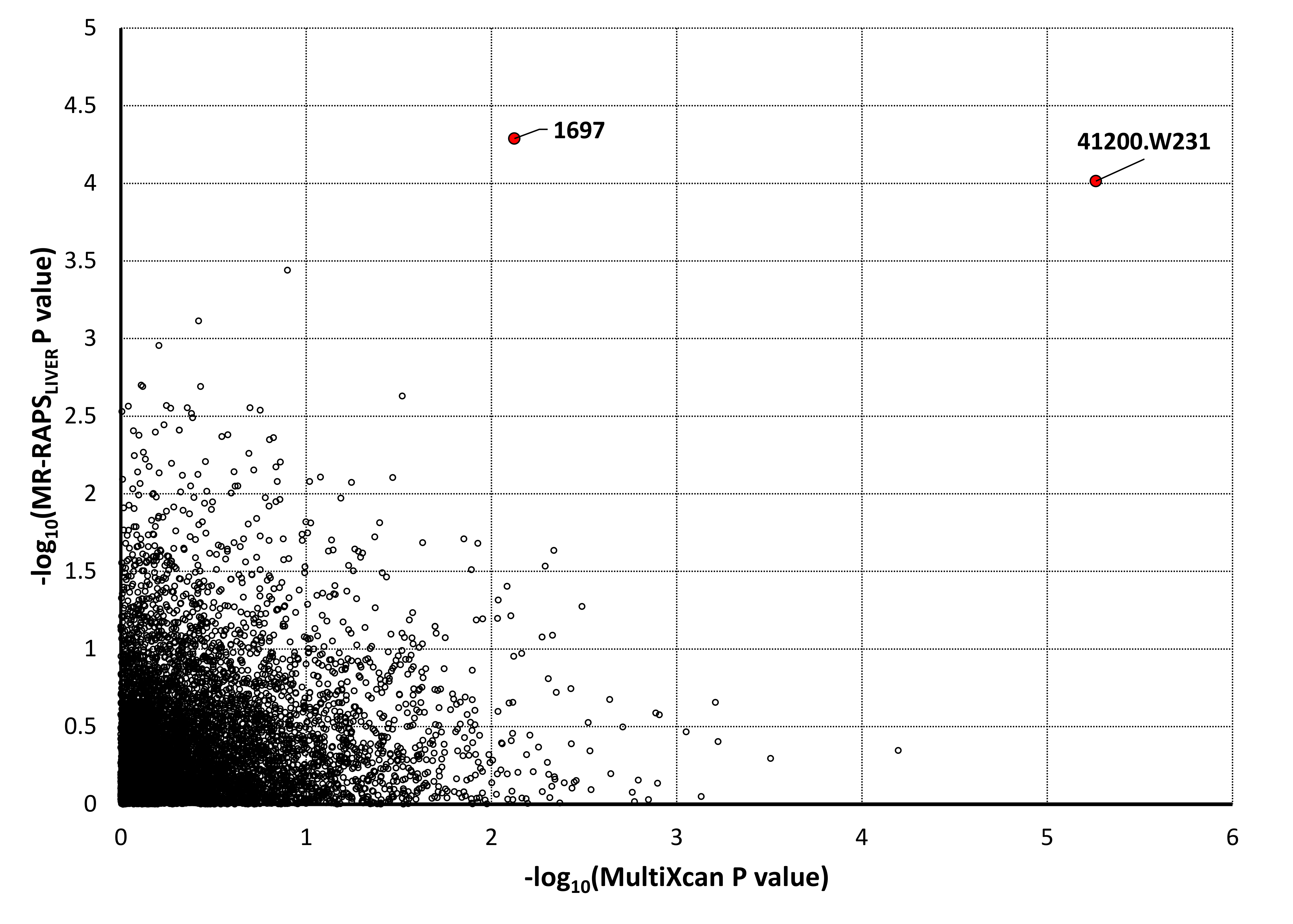

**Supplementary Figure 2**: Concordance between MultiXcan and MR-RAPS_liver_ associations across the 7,149 traits assessed in UK Biobank cohort.

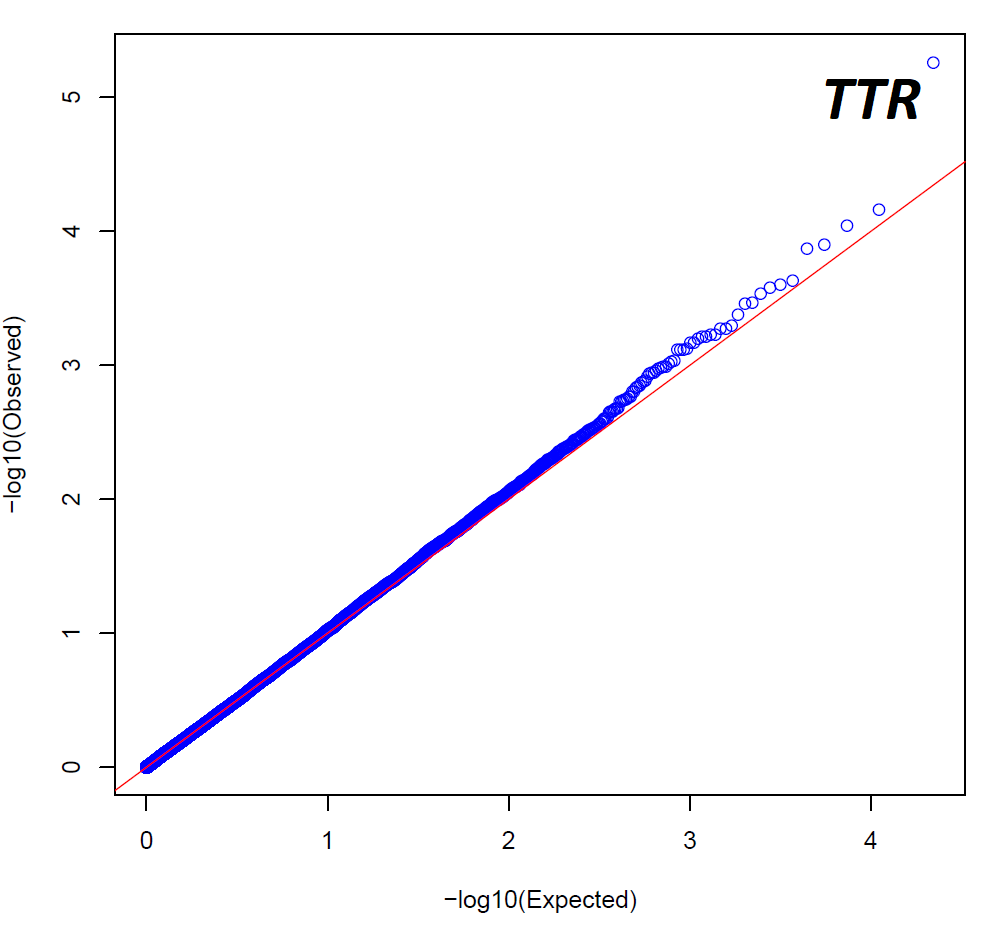

**Supplementary Figure 3**: QQ plot (λ_gc_=0.996) of the transcriptome-wide association statics related to the phenotype 41200-W231.
